# Surge and decline of typhoid fever before and after a typhoid conjugate vaccine trial: a decade of blood culture surveillance in Asante Akim North, Agogo, Ghana

**DOI:** 10.64898/2026.11.17.26368362

**Authors:** Sampson Twumasi-Ankrah, Michael Owusu-Ansah, Seidu Amenyaglo, Caleb Osei-Wusu Sarfo, Portia Okyere Boakye, Michael Owusu, Kennedy Gyau Boahen, Francis Opoku Agyapong, Samuel Nkansah-Darko, Akua Serwaa Boafo, Adwoa Safowaa, Seung Eun Kyung, Birkneh Tilahun Tadesse, Asma Aziz, Florian Marks, Yaw Adu-Sarkodie, Ellis Owusu-Dabo

## Abstract

Long surveillance series that follow typhoid fever through a surge and its aftermath are rare in Africa. Here, we describe ten years of blood culture surveillance in Agogo, Ghana; the period included a surge in 2017 and a cluster randomised trial of a typhoid conjugate vaccine in 2021–2022. We analysed 20,035 blood cultures from febrile patients enrolled between May 2016 and December 2025 at Asante Akim North, Agogo. Periods were identified by change point detection, and changes in positivity were estimated using segmented binomial regression. With the trial’s household census as the denominator, we estimated detected incidence (culture confirmed cases detected per 100,000 person-years) among residents of the 15 trial communities. In total, 82 cultures (0.41%) grew *Salmonella enterica* serovar Typhi. Positivity rose from 2.77% (95% confidence interval 1.48–4.69) to 8.41% (6.05–11.31) during a surge between February and December 2017 and fell to 0.16% (0.11–0.22) from 2018. Thirty of 39 surge cases were children aged 5–14 years. Among residents, the detected incidence was 65.3 (45.3–91.3) per 100,000 person-years during the surge and 5.4 (2.5–10.2) in the post-surge years before vaccination. It was 5.9 (3.5–9.4) per 100,000 person-years after a campaign that vaccinated 87% of eligible children, while testing per head rose more than eight-fold. After adjustment for care seeking and blood culture sensitivity, incidence in 2016–2019 was 141 (104–189) per 100,000 person-years. Typhoid in Agogo fell sharply after 2017, three years before vaccination. Most of the later fall in test positivity reflects more testing rather than fewer cases. Thus, we recommend continued surveillance with population denominators.

## Introduction

Typhoid fever, caused by *Salmonella enterica* serovar Typhi (*S*. Typhi), was estimated to cause 10.9 million illnesses and 116,800 deaths worldwide in 2017 [1]. Recent estimates for Ghana are about 20,600 cases and 330 deaths a year, three-quarters of them in children younger than 15 years [2]. In Africa, population-based surveillance in the Typhoid Fever Surveillance in Africa Program (TSAP) and the Severe Typhoid in Africa programme (SETA) reported incidence rates above 100 per 100,000 person-years at several sites, including Agogo in the Ashanti Region of Ghana [3, 4]. However, incidence varied markedly over time and between places [5]. Long series from the same site are rare, as each programme covered only a few years.

Typhoid also occurs as surges, when a contaminated source or a new strain reaches a susceptible population. In Blantyre, Malawi, multidrug-resistant typhoid rose rapidly from 2011 [6], whereas in South Africa, typhoid declined over three decades [7]. What follows a surge is relevant to vaccine policy. Transmission models predict that, once transmission falls, susceptible children accumulate through births and waning immunity, so that incidence can rebound five to fifteen years later [8–10]. Few African surveillance series are long enough to follow a population through a surge and its aftermath. In a PubMed search through October 2026, we found no study that followed one population through a typhoid surge and a typhoid conjugate vaccine trial with census denominators.

Agogo offers an unusual opportunity to do so. Blood culture surveillance at the district hospital has been conducted under a stable protocol since 2016 [11]. In August 2021, the Evaluation of Typhoid Conjugate Vaccine Effectiveness in Ghana (TyVEGHA) trial began vaccinating children aged 9 months to 15 years in 80 clusters of the surrounding municipality, using either the Vi-tetanus toxoid conjugate vaccine (Vi-TT) or a meningococcal control vaccine [12, 13]. The trial enumerated every household in its 15 communities and updated the census three times. Thus, it provides population denominators that hospital surveillance usually lacks. With them, detected incidence, and not only test positivity, can be estimated, and changes in testing can be separated from changes in cases. In context, Ghana has not yet introduced a typhoid conjugate vaccine into its national immunisation programme; in early 2026 the Ghana Health Service announced vaccination of food handlers [2, 14]. As Ghana considers national introduction, it is important to know whether a fall in test positivity reflects fewer cases or more testing. Short surveillance windows, on which many incidence estimates and decisions on vaccine introduction are based, may capture an unusual period.

Here, we describe what is, to our knowledge, the longest blood culture series of typhoid from one African site with population denominators. We had three aims: (1) to identify the surge and the periods around it with methods suited to count data; (2) to estimate changes in test positivity and in detected incidence, taking account of changes in testing; and (3) to describe who was affected, by age and sex.

## Materials and methods

### Ethics statement

The Committee on Human Research, Publications and Ethics of Kwame Nkrumah University of Science and Technology approved the SETA surveillance protocol (CHPRE/AP/219/16). The Ghana Health Service Ethics Review Committee (GHS-ERC) approved the TyVEGHA trial (GHS-ERC:010/04/21), and the Food and Drugs Authority (FDA) approved it (FDA/CT/219 (2 ext 1). Written informed consent was obtained from all participants, or from the parents or guardians of minors; children aged 12 years and older gave written assent.

### Setting and surveillance

Agogo Presbyterian Hospital is the referral hospital of Asante Akim North Municipality, which had 85,788 residents at the 2021 national census [15]. Under SETA (2016–2019) and its extension, the Severe Typhoid in Africa Plus programme (SETA Plus; 2019 onwards), febrile patients of any age were offered enrolment and a blood culture [11, 16]; fever was defined as ≥38.0°C tympanic or ≥37.5°C axillary, or reported fever for at least 3 of the previous 7 days. The protocol was amended several times (SETA Plus versions 3.0 in November 2019, 4.0 in February 2021 and 7.0 in October 2024). However, the amendments concerned follow-up, additional study components and administration. The recruiting hospital, the case definition and eligibility criteria, blood-culture collection, and the laboratory and its identification methods were the same throughout 2016–2025. Blood was incubated in a BACTEC FX40 system. *S*. Typhi was identified by colony morphology, Analytical Profile Index (API) 20E and agglutination. Cultures that grew only organisms regarded as contaminants (coagulase-negative staphylococci, *Bacillus, Micrococcus, Corynebacterium* or viridans streptococci) were classified as negative for *S*. Typhi. Patients enrolled on a history of fever alone were included in both the numerator and the denominator of positivity, so temperature at presentation was normal in some cases. We included all cultures recorded between 3 May 2016 and 12 December 2025, and the reported place of residence for each patient was matched to the 15 TyVEGHA communities. Cultures were available in 106 of the 116 calendar months; none were recorded in May–August 2019, October 2019–February 2020 and April 2020, during the transition from SETA to SETA Plus. These months were treated as missing rather than as months without cases: the change-point analysis was applied to the 106 observed months in order, and the regression models used calendar time.

### Population denominators

Population denominators were derived from the TyVEGHA household census of the 15 trial communities (2019–2020), with later updates through December 2025. Before the baseline census, the population was back-projected at 1.98% per year; this is the average annual growth of Asante Akim North between the 2010 (69,186) and 2021 (85,788) national censuses [15, 17], and the exact census dates give 2.02%. However, the growth rate at the community level is not known. Thus, we repeated the incidence analyses with rates of 1.0%, 2.5%, 3.0% and 4.3% per year.

### Vaccination records

Between 23 August 2021 and 15 March 2022, the trial vaccinated 20,050 of the 22,973 eligible children with one of the two study vaccines, representing 87.3% coverage [12, 13]. Allocation was by cluster and remains blinded. However, vaccine accountability records showed that 10,035 children received Vi-TT (43.7% of those eligible). The effectiveness analysis of the trial is not yet available.

### Statistical analysis

#### Periods

Cultures and *S*. Typhi isolates were aggregated by calendar month. Periods were identified with the Pruned Exact Linear Time (PELT) algorithm [18] using a binomial cost, so that months with few cultures carried less weight. The primary analysis used a Bayesian information criterion (BIC) penalty and a minimum segment of 6 months. We repeated it with beta-binomial and normal likelihoods, five penalties, and minimum segment lengths of 3, 6, and 12 months. We also estimated the uncertainty of each boundary with a parametric bootstrap (500 replicates).

#### Positivity, months without cases and detected incidence

Changes in positivity were estimated using segmented binomial regression of monthly isolates from cultures [19, 20], with level and slope terms at the period boundaries and mean daily rainfall as a covariate. Rainfall was the Climate Hazards Group Infrared Precipitation with Stations (CHIRPS) daily estimate from Google Earth Engine [21], averaged across the 15 trial communities and over the recruitment days in each month. The model was checked for overdispersion and residual autocorrelation, and it was refitted with beta-binomial, observation-level random-effect and first-order autoregressive extensions [22]. In sensitivity analyses, we adjusted for the age mix of tested patients, added a vaccine-era indicator, and used alternative boundaries. Months without a case were analysed using a hurdle model [23, 24] (S1 Appendix, Tables S4 and S8), with a logistic part for whether a month had any case and a zero-truncated Poisson part for the number of cases per culture. We also checked whether months without cases exceeded the number expected under binomial sampling given the number of cultures. Because every surge month had a case, pre-surge and surge months were combined in the occurrence component.

Detected incidence was defined as the number of culture confirmed cases detected among residents of the 15 communities divided by person-time, summed over months with surveillance; it is reported as detected cases per 100,000 person-years. It counts only people who came to the hospital, were tested and had a positive culture. It is therefore lower than true incidence, by a factor that depends on care seeking, recruitment and the sensitivity of a single blood culture, none of which we measured. Thus, we used it to compare periods within the same system rather than to estimate burden. Periods were compared with negative binomial regression with a person-time offset, and age-specific incidence used the baseline census age distribution. To compare our estimates with SETA, the calculation was repeated for SETA’s reporting window from May 2016 to April 2019 [25].

To approximate the true incidence, we divided the detected incidence by the product of two probabilities: the probability that a febrile resident first sought care at the hospital and the probability that a single blood culture detected *S*. Typhi. The first was taken from the third update of the TyVEGHA census (October–December 2025), which asked whether any household member had had fever for at least two days in the previous three months and where treatment was first sought. Culture sensitivity was modelled from the blood volumes recorded in the SETA database for 2016–2019 (0.51 at 2 mL to 0.65 at 10 mL) and was 34% lower in patients who had taken antibiotics [26]. Uncertainty was propagated by Monte Carlo simulation (S1 Appendix, section 1.5). We also divided by the proportion of eligible febrile residents seen at the hospital who were enrolled, taken from the SETA Plus screening and enrolment logs for Agogo (January 2022 to February 2026), and assumed that the same proportion applied before 2022. Contamination of blood cultures at Agogo was taken from the laboratory contamination log (October 2021 to December 2025). To check whether detection changed over time, we tabulated, by year, the self-reported use of antibiotics before enrolment and the number of days with fever before the visit.

#### Age and sex

Age and sex were compared across periods with exact binomial confidence intervals and Fisher’s exact test. These comparisons are descriptive, and we did not adjust for multiple testing. Analyses were conducted in R version 4.3.3 [27]. The study is reported in accordance with the Strengthening the Reporting of Observational Studies in Epidemiology (STROBE) guidelines and the REporting of studies Conducted using Observational Routinely-collected health Data (RECORD) extension for studies using routinely collected health data [28].

## Results

### Surveillance

Between May 2016 and December 2025, 20,035 cultures were performed, and 82 (0.41%) grew *S*. Typhi. Testing changed considerably over the decade (Fig 1B). About 40–50 cultures per month were collected until 2018, almost none in late 2019 and early 2020, and 300–650 per month from late 2022, when the enhanced surveillance of the trial was in place. Overall, 17,566 cultures (87.7%) and 68 cases came from residents of the trial communities.

**Fig 1.**
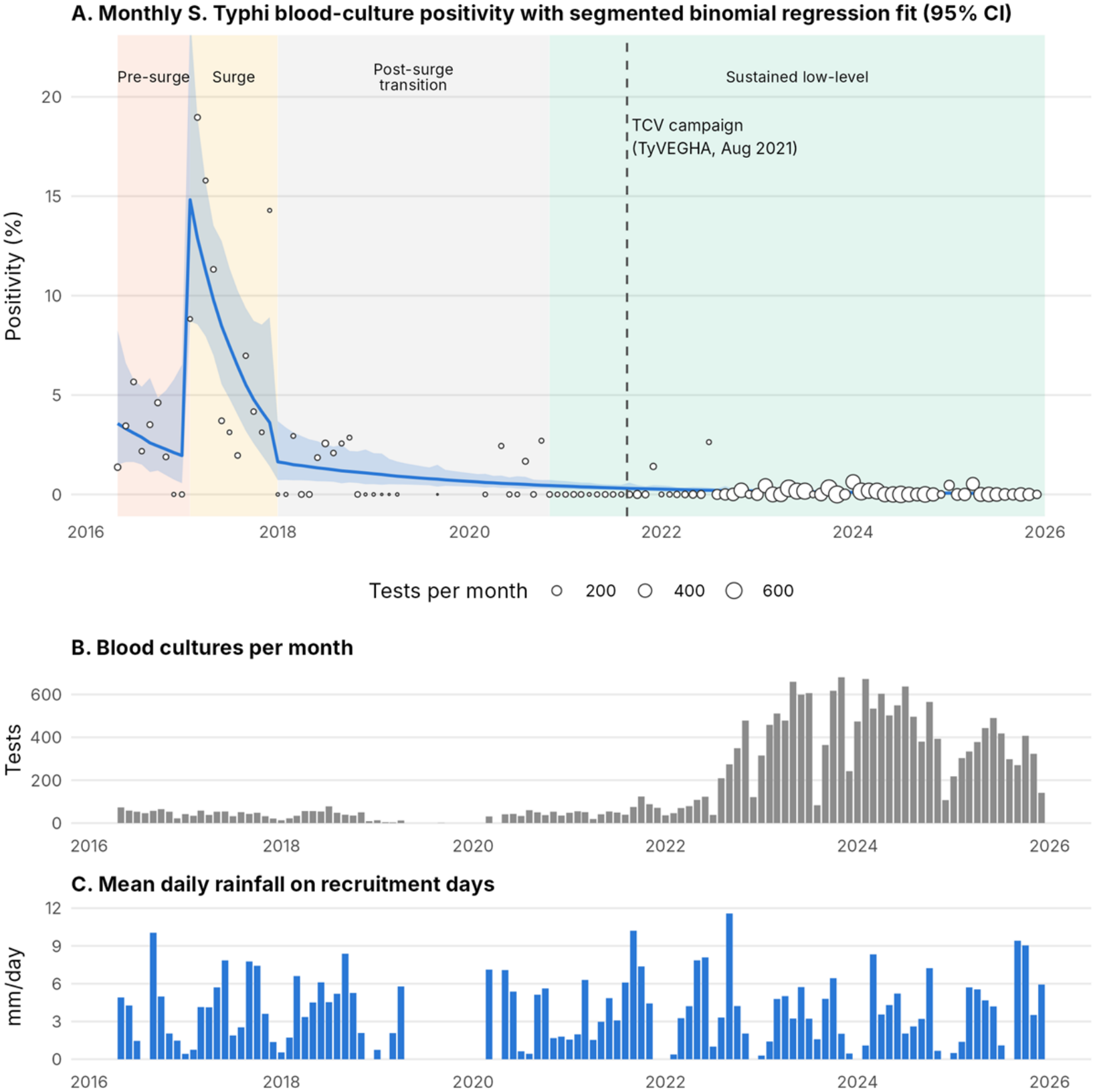
Blood-culture positivity for Salmonella Typhi in Agogo, Ghana, May 2016 to December 2025. (A) Monthly positivity (circles, sized by number of cultures) with the fitted segmented binomial regression and 95% confidence interval. Shading marks the periods from change-point detection; the dashed line marks the start of the TyVEGHA vaccination campaign. (B) Blood cultures per month. (C) Mean daily rainfall on recruitment days.

### Periods

Binomial change point detection identified new segments starting in February 2017, January 2018, November 2020 and May 2024 (Fig 1A). We used the first two to define a pre-surge period (May 2016 to January 2017), a surge period (February to December 2017), and a post-surge period (January 2018 onwards) (Table 1). The end of the surge was the most stable boundary; it fell within a month of January 2018 in 14 of 15 binomial configurations and in 83% of bootstrap replicates. However, the onset was less stable (in 75% of replicates), and the later boundaries appeared in fewer than half of replicates (S1 Appendix, Figure S1). The two later boundaries reflect changes in testing more than in cases. From January 2018 to October 2020 there were 34 cultures a month and positivity was 1.22% (95% confidence interval [CI] 0.58–2.23%); from November 2020 to April 2024, 253 cultures a month and 0.16% (0.09–0.26%); and from May 2024, 383 cultures a month and 0.04% (0.01–0.11%). Detected incidence among residents over the same intervals was 7.7, 6.5 and 2.4 per 100,000 person-years. We therefore treated the post-surge period as a single positivity segment and divided it at the start of vaccination for incidence.

**Table 1.**
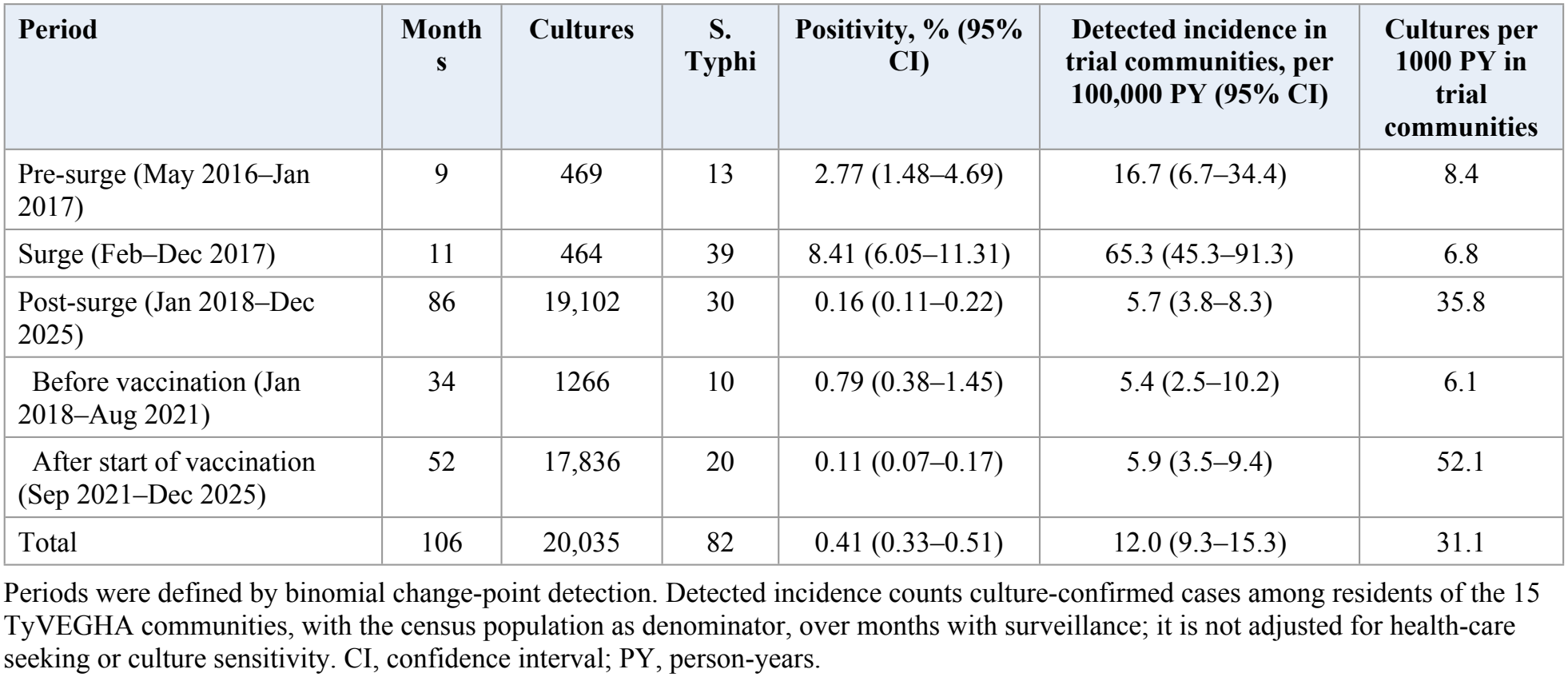
Blood culture positivity and detected incidence of typhoid by period, Agogo, 2016–2025.

| **Period** | **Month s** | **Cultures** | **S. Typhi** | **Positivity, % (95% CI)** | **Detected incidence in trial communities, per 100,000 PY (95% CI)** | **Cultures per 1000 PY in trial communities** |
|---|---|---|---|---|---|---|
| Pre-surge (May 2016–Jan 2017) | 9 | 469 | 13 | 2.77 (1.48–4.69) | 16.7 (6.7–34.4) | 8.4 |
| Surge (Feb–Dec 2017) | 11 | 464 | 39 | 8.41 (6.05–11.31) | 65.3 (45.3–91.3) | 6.8 |
| Post-surge (Jan 2018–Dec 2025) | 86 | 19,102 | 30 | 0.16 (0.11–0.22) | 5.7 (3.8–8.3) | 35.8 |
| Before vaccination (Jan 2018–Aug 2021) | 34 | 1266 | 10 | 0.79 (0.38–1.45) | 5.4 (2.5–10.2) | 6.1 |
| After start of vaccination (Sep 2021–Dec 2025) | 52 | 17,836 | 20 | 0.11 (0.07–0.17) | 5.9 (3.5–9.4) | 52.1 |
| Total | 106 | 20,035 | 82 | 0.41 (0.33–0.51) | 12.0 (9.3–15.3) | 31.1 |

Periods were defined by binomial change-point detection. Detected incidence counts culture-confirmed cases among residents of the 15 TyVEGHA communities, with the census population as denominator, over months with surveillance; it is not adjusted for health-care seeking or culture sensitivity. CI, confidence interval; PY, person-years.

### Changes in positivity

Observed positivity peaked at 18.97% in March 2017 (11 of 58 cultures; exact 95% CI 9.9–31.4%). In the segmented model, the start of the surge was associated with a ninefold increase in the odds of a positive culture. Positivity then declined steadily, by 37% per year in the odds after the surge (Table 2). The data were neither overdispersed (Pearson dispersion 1.02) nor autocorrelated (Ljung–Box p = 0.32), and the three extended models gave the same estimates. The surge effect held in all sensitivity analyses. However, it halved to an odds ratio of 4.39 (95% CI, 1.03–18.61) when the onset was set to January 2017, a month with no positive culture (S1 Appendix, Tables S2 and S3).

**Table 2.** Changes in typhoid positivity and detected incidence between periods.

| **Model and term** | **Estimate (95% CI)** | **p** |
|---|---|---|
| *Segmented binomial regression of monthly positivity* |  |  |
| Level change at start of surge (Feb 2017), odds ratio | 9.47 (2.05–43.79) | 0.004 |
| Trend during surge, odds ratio per year | 0.16 (0.04–0.69) |  |
| Level change at end of surge (Jan 2018), odds ratio | 0.51 (0.15–1.71) | 0.27 |
| Trend after surge, odds ratio per year | 0.63 (0.54–0.74) |  |
| Rainfall, odds ratio per 5 mm per day | 0.97 (0.61–1.53) | 0.90 |
| *Hurdle model of monthly counts* |  |  |
| Any case in month, post-surge vs earlier, odds ratio<sup>a</sup> | 0.029 (0.006–0.144) | <0.001 |
| Cases per culture in months with cases, surge vs pre-surge, rate ratio | 3.41 (1.51–7.71) | 0.003 |
| Cases per culture in months with cases, post-surge vs pre-surge, rate ratio | 0.073 (0.026–0.202) | <0.001 |
| *Detected incidence in trial communities, negative binomial regression* |  |  |
| Surge vs pre-surge, rate ratio | 3.92 (1.35–11.39) | 0.012 |
| Post-surge before vaccination vs pre-surge, rate ratio | 0.32 (0.10–1.01) | 0.052 |
| Post-surge after start of vaccination vs pre-surge, rate ratio | 0.35 (0.13–0.99) | 0.048 |

Beta-binomial, observation-level random-effect and autoregressive versions of the regression gave the same estimates (S1 Appendix, Table S2). <sup>a</sup> Adjusted for log number of cultures and rainfall; pre-surge and surge months were combined because every surge month had a case. CI, confidence interval.

65 of 106 months had no positive culture: 2 of 9 pre-surge months, none of 11 surge months and 63 of 86 post-surge months. A binomial model with the observed numbers of cultures predicted 65.9 such months (2.3, 0.4 and 63.9 by period); thus, there was no excess of zeros. In the hurdle model, the odds of a month having any case were 97% lower after the surge, after adjustment for the number of cultures. In months with cases, the number of cases per culture was 93% lower (Table 2).

### Detected incidence

Among residents of the clinical trial communities, detected incidence quadrupled during the surge and then fell to about a third of its pre-surge level (Table 1, Fig 2). The fall had occurred by 2018–2020, three years before vaccination, when testing per head was similar to that before the surge. After the campaign began, testing per head rose more than eight-fold; however, detected incidence did not change appreciably (5.9 vs 5.4 per 100,000 person-years). Positivity therefore fell much further than incidence. In other words, most of the drop in positivity after 2021 reflects more testing rather than fewer cases. These results were insensitive to the growth rate used for back-projection (S1 Appendix, Table S7).

**Fig 2.**
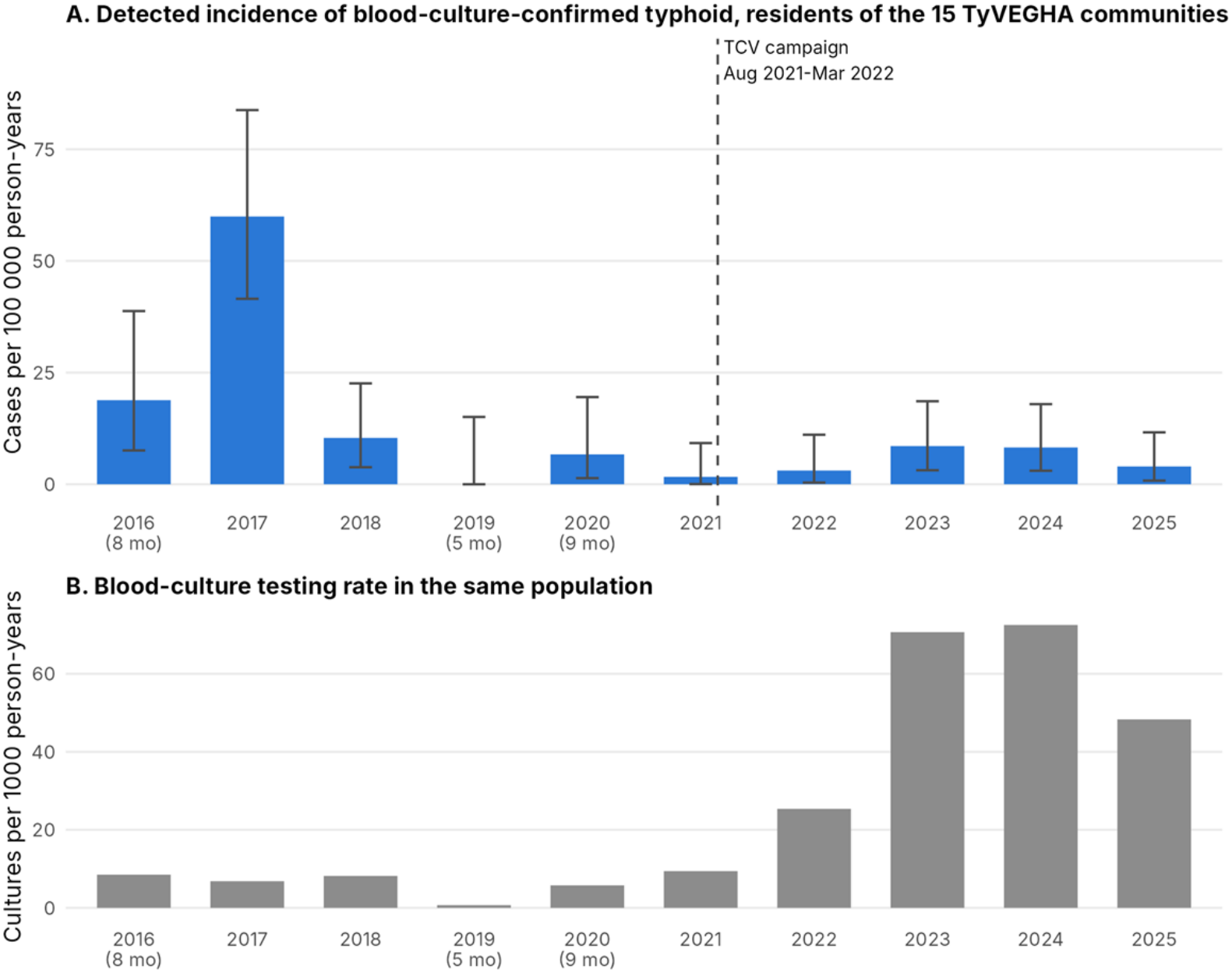
Detected incidence and testing among residents of the 15 TyVEGHA trial communities. (A) Culture-confirmed typhoid per 100,000 person-years with exact 95% confidence intervals, using the census population as denominator; years with incomplete surveillance are labelled with the number of months covered. (B) Blood cultures per 1000 person-years.

Over the SETA reporting window (May 2016 to April 2019), 47 cases were detected among residents of the trial communities in 171,277 person-years, a crude detected incidence of 27.4 (95% CI 20.2–36.5) per 100,000 person-years (S1 Appendix, Table S9). For the same window, SETA reported 60 cases at Agogo, with a crude incidence of 10 per 100,000 and an incidence of 112 (84–164) per 100,000 after adjustment for health-care seeking and blood culture sensitivity [4, 25].

During the surge, the detected incidence among children aged 5–14 years was 175 per 100,000 person-years (95% CI, 113–259), compared with 93 per 100,000 person-years (30–218) at 0–4 years and 12 per 100,000 person-years (3–32) at 15 years and older (Table 3). In children aged 5–14 years, it was 15 (6–31) after the surge but before vaccination, and 10 (4–19) after the campaign began.

**Table 3.** Typhoid positivity and detected incidence by age group and period.

| Age group | Pre-surge | Surge | Post-surge |
|---|---|---|---|
| *Positivity, positive/tested, % (95% CI)* |  |  |  |
| <5 years | 4/171, 2.3 (0.6–5.9) | 5/168, 3.0 (1.0–6.8) | 5/7241, 0.07 (0.02–0.16) |
| 5–14 years | 7/102, 6.9 (2.8–13.6) | 30/137, 21.9 (15.3–29.8) | 18/5349, 0.34 (0.20–0.53) |
| 15–44 years | 2/129, 1.6 (0.2–5.5) | 4/104, 3.8 (1.1–9.6) | 7/4668, 0.15 (0.06–0.31) |
| ≥45 years | 0/67, 0 (0–5.4) | 0/55, 0 (0–6.5) | 0/1844, 0 (0–0.20) |
| *Detected incidence in trial communities, per 100,000 PY (95% CI)* |  |  |  |
| 0–4 years | 46 (6–167) | 93 (30–218) | 10 (3–24)<sup>a</sup> |
| 5–14 years | 26 (5–76) | 175 (113–259) | 12 (6–19)<sup>a</sup> |
| ≥15 years | 8 (1–28) | 12 (3–32) | 2.4 (1.0–4.9)<sup>a</sup> |

<sup>a</sup> Whole post-surge period; before vaccination: 5–14 years 15 (6–31), ≥15 years 1.9 (0.2–6.9), 0–4 years no cases; after start of vaccination: 0–4 years 16 (5–37), 5–14 years 10 (4–19), ≥15 years 2.6 (0.9–6.2). Age-specific person-time used the baseline census age distribution (0–4 years 10.3%, 5–14 years 27.4%, ≥15 years 62.3%). CI, confidence interval; PY, person-years.

### Age and sex

Thirty of 39 surge cases (77%) were among children aged 5–14 years, with positivity reaching 21.9% (95% CI 15.3–29.8). Across the pre-surge and surge periods, the odds of a positive culture at 5–14 years were 6.7 times those at younger than 5 years (3.2–14.2) (Table 3, Fig 3). No case occurred in patients aged 45 years or older. Positivity was higher in males than in females before the surge (4.39% vs 1.24%; p = 0.049), but not during it (8.94% vs 7.86%; p = 0.74) or after it (0.21% vs 0.11%; p = 0.10).

**Fig 3.**
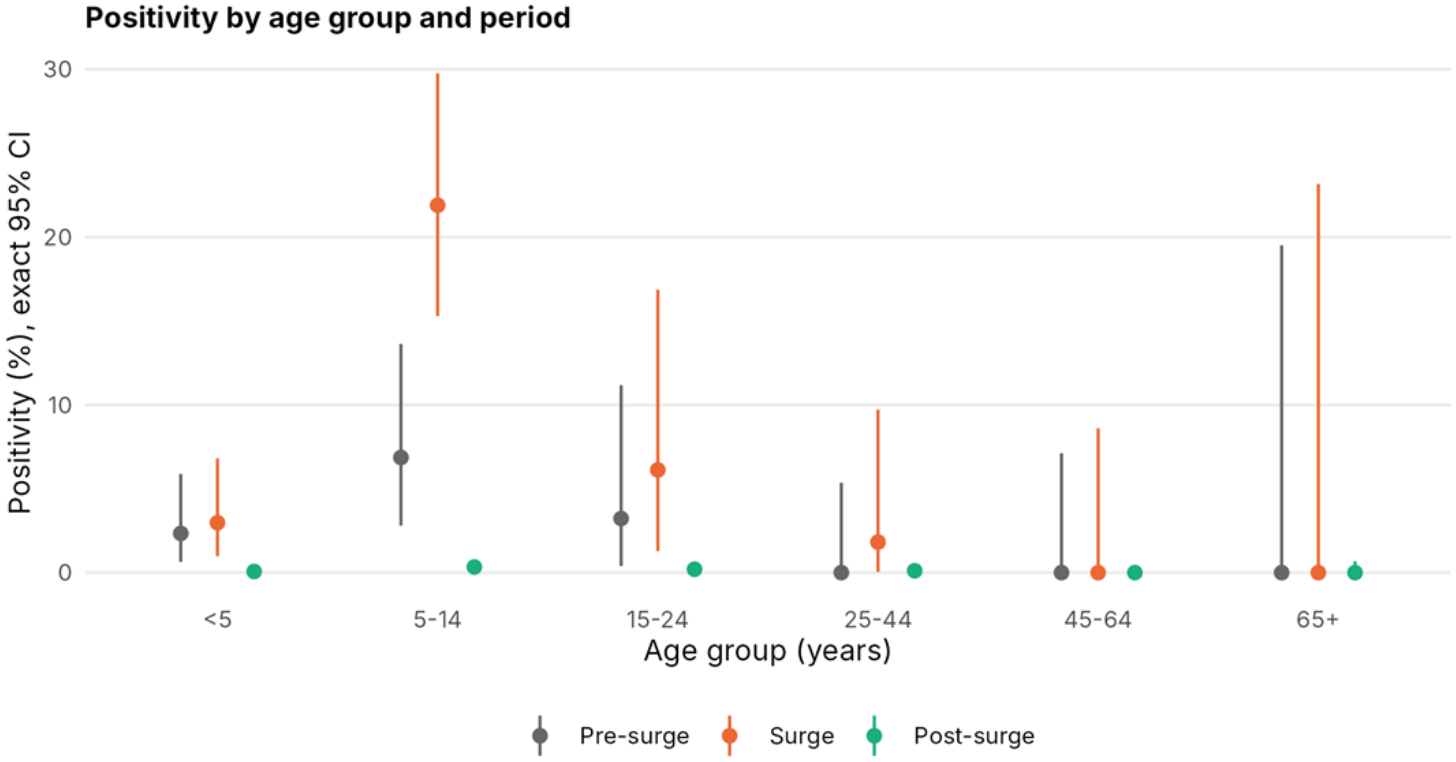
Typhoid positivity by age group and period. Points show positivity and bars exact 95% confidence intervals.

### Estimated true incidence

Detection did not appear to worsen over time. Self-reported antibiotic use before enrolment fell from 10.8% of patients before the surge to 0.7% in the vaccine era. The share of those who had had a fever for more than five days fell from 16–19% in 2016–2018 to 8–9% in 2023–2025 (S1 Appendix, Table S10). In the Agogo laboratory contamination log, 3.4% of cultures in 2022, 3.3% in 2023, 5.5% in 2024 and 4.2% in 2025 grew only contaminants, mainly coagulase-negative staphylococci; contamination was not logged before October 2021. Both changes should make a culture more likely to detect *S*. Typhi. In the third census update survey, 35.5% (95% CI 33.8–37.1%) of recent febrile illnesses were first treated at a hospital (48.6% in Agogo town and 22.9% in the other communities; S1 Appendix, Table S11). The SETA Plus screening logs for Agogo (January 2022 to February 2026) recorded 18,250 eligible febrile residents, of whom 17,918 (98.2%, 95% CI 98.0–98.4%) were enrolled (S1 Appendix, Table S12). Together, these figures imply that at least five cases occurred for every one detected (multiplier 5.1, 95% uncertainty interval 4.7–5.7). Estimated true incidence over the SETA window was 141 (104–189) per 100,000 person-years, close to the SETA adjusted estimate of 112 (84–164) [25]. It was 337 during the surge and about 28–30 per 100,000 person-years after it (S1 Appendix, Table S13).

## Discussion

Over ten years in Agogo, typhoid cases rose sharply in early 2017, mostly among school-aged children, and then fell to low levels that have persisted through the end of 2025. Two findings stand out. First, the decline was largely real; detected incidence among residents of the trial communities fell by about two-thirds by 2018–2020, when testing was no more intense than before the surge. Second, the much steeper fall in test positivity after 2021 was largely produced by an eight-fold increase in testing per head, not by a further fall in cases.

Both findings depend on population denominators, which hospital surveillance rarely has. Without the TyVEGHA census, we would have described a 25-fold fall in positivity from pre-surge levels and might have credited much of it to the vaccine. With the census, the detected incidence after the campaign (5.9 per 100,000 person-years) was close to that in the three years before it (5.4 per 100,000 person-years). However, this comparison cannot estimate vaccine effectiveness. Fewer than half of eligible children received Vi-TT, incidence was already low, and the numbers are small. In addition, wider testing may have found milder cases that would previously have been missed, thereby masking a real reduction. Thus, the cluster level comparison of the trial is the appropriate source for that estimate.

Two further checks support this interpretation. In recent years, patients presented earlier and reported fewer antibiotic exposures, both of which should make culture more sensitive; detection is therefore unlikely to have worsened. The screening logs show that almost every eligible resident seen at the hospital was enrolled from 2022 onwards, so recruitment did not limit detection in the vaccine era. In addition, after adjustment for care seeking and culture sensitivity, incidence in 2016–2019 agreed with the independent SETA estimate. This implies that at least five cases occurred in these communities for every one we detected. About 30 cases per 100,000 person-years after the surge is low; however, it is not elimination.

The coronavirus disease 2019 (COVID-19) pandemic overlapped the post-surge period. Ghana closed schools on 15 March 2020, imposed a three-week partial lockdown in Accra and Kumasi from 30 March 2020, and fully reopened schools in January 2021. These measures cannot explain the fall in typhoid, which was complete by early 2018, two years before the pandemic, and positivity and detected incidence stayed low after schools reopened. The pandemic did coincide with the transition to SETA Plus, with no cultures in most of the first half of 2020, so typhoid during this period is the least well measured part of the series.

The reasons for the fall in typhoid after 2017 remain uncertain. Possible explanations include the end of a point-source exposure, changes in water, sanitation, or food handling after the outbreak, or the disappearance of an outbreak strain. Wastewater sampling in Agogo found *S*. Typhi in 44% of samples, including in months without clinical cases [29]. Thus, the organism has not disappeared.

The concentration of the surge among children aged 5–14 years, with a detected incidence of 175 per 100,000 person-years, agrees with SETA, TSAP and global burden data [1, 3, 4], and with models in which school-aged children drive transmission [8]. Ghana’s current plan targets food handlers [14]. However, our data suggest that children should also be included, as the World Health Organization (WHO) recommends for catch-up campaigns up to 15 years of age [30].

This study has some limitations. It comes from one district, and comparisons between periods assume that ascertainment was similar over time; the stable protocol and the trends in presentation support this assumption but cannot guarantee it. The population before the baseline census was back-projected, although the results were insensitive to the assumed growth rate.

In conclusion, a typhoid surge among school-aged children in Agogo in 2017 was followed by a lasting decline that began before the conjugate vaccine trial. Changes in testing account for most of the later fall in positivity. Thus, we recommend sustained blood-culture surveillance with population denominators, recorded vaccine coverage, genomic data, and environmental monitoring, so that a resurgence can be detected early.

## Data Availability

Dataset are available at zenodo via: DOI 10.5281/zenodo.24200247

## Data availability

Monthly aggregated surveillance data are provided as S1 Data and all analysis code as S1 Code. Individual-level surveillance and census data contain identifying information and are available on reasonable request from the SETA Plus and TyVEGHA data-sharing committees. S1 Dataset are available at: DOI 10.5281/zenodo.24200247

## Acknowledgments

We thank the patients, families and communities of Asante Akim North, the staff of Agogo Presbyterian Hospital and the Kwame Nkrumah University of Science and Technology–International Vaccine Institute (KNUST-IVI) Collaborative Centre, and the TyVEGHA census and vaccination teams.

## Supporting information

S1 Appendix. Supplementary methods, tables and figure. Detailed methods for change-point detection, regression, hurdle models and the estimation of true incidence; Tables S1–S13, including sensitivity analyses for population growth and the hurdle count model; and Figure S1, robustness of the change points.

